# Unequal impact of structural health determinants and comorbidity on COVID-19 severity and lethality in older Mexican adults: Looking beyond chronological aging

**DOI:** 10.1101/2020.05.12.20098699

**Authors:** Omar Yaxmehen Bello-Chavolla, Armando González-Díaz, Neftali Eduardo Antonio-Villa, Carlos A. Fermín-Martínez, Alejandro Márquez-Salinas, Arsenio Vargas-Vázquez, Jessica Paola Bahena-López, Carmen García-Peña, Carlos A. Aguilar-Salinas, Luis Miguel Gutiérrez-Robledo

**Affiliations:** Division of Research, Instituto Nacional de Geriatría; Unidad de Investigación de Enfermedades Metabólicas, Instituto Nacional de Ciencias Médicas y Nutrición Salvador Zubirán; MD/PhD (PECEM), Faculty of Medicine, National Autonomous University of Mexico; Universidad Nacional Autónoma de México; Department of Endocrinolgy and Metabolism. Instituto Nacional de Ciencias Médicas y Nutrición Salvador Zubirán; Tecnologico de Monterrey, Escuela de Medicina y Ciencias de la Salud

**Keywords:** COVID-19, SARS-CoV-2, Human Aging, Inequality, Mortality, Mexico

## Abstract

**BACKGROUND:** COVID-19 has had a disproportionate impact on older adults. Mexico’s population is younger, yet COVID-19’s impact on older adults is comparable to countries with older population structures. Here, we aim to identify health and structural determinants that increase susceptibility to COVID-19 in older Mexican adults beyond chronological aging.

**METHODS:** We analyzed confirmed COVID-19 cases in older adults using data from the General Directorate of Epidemiology of Mexican Ministry of Health. We modeled risk factors for increased COVID-19 severity and mortality, using mixed models to incorporate multilevel data concerning healthcare access and marginalization. We also evaluated structural factors and comorbidity profiles compared to chronological age for improving COVID-19 mortality risk prediction.

**RESULTS:** We analyzed 7,029 confirmed SARS-CoV-2 cases in adults aged ≥60 years. Male sex, smoking, diabetes, and obesity were associated with pneumonia, hospitalization and ICU admission in older adults, CKD and COPD were associated with hospitalization. High social lag indexes and access to private care were predictors of COVID-19 severity and mortality. Age was not a predictor of COVID-19 severity in individuals without comorbidities and structural factors and comorbidities were better predictors of COVID-19 lethality and severity compared to chronological age. COVID-19 baseline lethality hazards were heterogeneously distributed across Mexican municipalities, particularly when comparing urban and rural areas.

**CONCLUSIONS:** Structural factors and comorbidity explain excess risk for COVID-19 severity and mortality over chronological age in older Mexican adults. Clinical decision-making related to COVID-19 should focus away from chronological aging onto more a comprehensive geriatric care approach.

## INTRODUCTION

The novel SARS-CoV-2 has disproportionately affected older adults. Notably, the impact of the COVID-19 pandemic has had larger repercussions in countries with older population structures; data from large outbreaks in China, Italy and, most recently, the United States has shown a remarkable impact of SARS-CoV-2 infections on older patients, with increased disease severity, adverse outcomes and increased mortality [1–3]. This susceptibility has been suggested to be attributable to deleterious features of aging, particularly in those with cardio-metabolic and respiratory comorbidities. Tissue-specific expression of the *ACE2* gene, which encodes the SARS-CoV-2 receptor, has been linked to specific immune signatures in males and older adults and its expression have been shown to be age-dependent [4,5]; moreover, immunosenescence and increased inflammatory responses related to the aging process might lead to increased infection risk and dysregulated immune response to SARS-CoV-2 in older adults [6]. Given these mechanisms which may increase susceptibility in older individuals, the role of comorbidities and social inequities, which condition an increased burden on functionality and dependence, has often been overlooked over chronological aging as significant predictors of COVID-19 severity and lethality. In Mexico, COVID-19 has shown similar patterns on its impact on older adults, despite an apparent demographic dividend by having, on average, a younger population [7]. Mexico’s younger population structure compared to European countries is nonetheless affected by increased rates of cardio-metabolic diseases, particularly type 2 diabetes, obesity and the metabolic syndrome, with most individuals reaching older age with a significant number of comorbidities [8,9]. Recent shifts on aging structures within Mexico have resulted in an increased number of older adults who continue working into old age, most of whom live in poverty, do not have adequate healthcare access, and are dependent on the working-age population [7]. The combination of unequal structural factors, an aging population with increased risk for chronic diseases, and the co-existence of infectious diseases position Mexican older adults as a particularly susceptible population during the COVID-19 pandemic [10]. Given that most studies have linked an increased risk of COVID-19 with chronological age, many resource allocation decisions have been made purely on this basis, which represents an issue for geriatric care in the patient population facing the highest disease burden [11]. To address these gaps, we investigated health and structural determinants which could contribute to increased COVID-19 severity and lethality beyond chronological age in older Mexican adults to better characterize the impact of COVID-19 in older populations.

## METHODS

### Data sources

We analyzed data collected by the General Directorate of Epidemiology of the Mexican Ministry of Health, which is an open-source dataset comprising daily updated suspected COVID-19 cases that have been tested for SARS-CoV-2 and were certified by the National Institute for Diagnosis and Epidemiological Referral [12]. Data to estimate population rates at different ages and population density was extracted from 2020 population projections obtained by the Mexican National Population Council (CONAPO). The 2015 social lag index, which is a composite of several factors which are measured to estimate social disadvantage and structural inequality based on population census data, was obtained from the Mexican National Evaluation Council (CONEVAL); the social lag index is a principal component score which comprises percentages of literacy, access to basic education, healthcare services, living conditions including drainage, dirt floor, access to water, electricity and electrical appliances for each Mexican municipality [13]. Data on available hospital beds at the end of 2019 were extracted from the Mexican Ministry of Health and were standardized to projected 2020 population figures from CONAPO [14]. For this work we considered all SARS-CoV-2 positive cases and, particularly, cases in individuals aged ≥60 years.

### Definitions of COVID-19 cases, predictors, and outcomes

Suspected COVID-19 cases were defined as an individual of any age whom in the last 7 days has presented ≥2 of the following: cough, fever, or headache, accompanied by either dyspnea, arthralgias, myalgias, sore throat, rhinorrhea, conjunctivitis or chest pain. Amongst suspected cases, the Ministry of Health establishes two protocols for case confirmation: 1) SARS-CoV-2 testing is done widespread for suspected COVID-19 cases with severe acute respiratory infection and signs of breathing difficulty or deaths with suspected COVID-19, 2) for all other cases, a sentinel surveillance model is being utilized, whereby 475 health facilities which comprise a nationally representative sample evaluate ~10% of mild outpatient cases to provide estimates of confirmed mild cases [15]. Demographic and health data are collected and uploaded to the epidemiologic surveillance database by personnel from each corresponding healthcare facility. Available variables include age, sex, nationality, state and municipality where the case was detected, immigration status as well as identification of individuals who speak indigenous languages from Mexico. Health information includes the status of diabetes, obesity, chronic obstructive pulmonary disease (COPD), immunosuppression, pregnancy, arterial hypertension, cardiovascular disease, chronic kidney disease (CKD), and asthma. Date of symptom onset, hospital admission, and death are available for all cases as an outpatient or hospitalized status, information regarding the diagnosis of pneumonia, ICU admission, and whether the patient required invasive mechanical ventilation.

### Statistical analysis

#### Population-based statistics

We estimated age-specific incidence and mortality rates by standardizing cases per 1-year increment to the corresponding projected population sizes and normalized it to reflect a rate per 100,000 inhabitants. Incidence and mortality rates were plotted against age and smoothed splines were fitted to model trends according to age profiles. Categorical variables were compared using the chi-squared statistic and stratified according to age.

#### COVID-19 outcome models

Given the large structural inequalities in healthcare access across Mexico, we fitted mixed-effects models which considered municipality of case occurrence as a random effect to control for potential inequalities in healthcare access, aging structures, and unequal regional evolution of the epidemic across regions. These models would also allow to include multilevel data, to increase the precision of outcome estimates considering structural factors for each municipality. Models were fitted using mixed-effects logistic regression using the municipality of case occurrence as a random effect; p-values and confidence intervals were estimated using the Laplace approximation within the *lme4* and *lmerTest* R packages.

#### Risk of COVID-19 lethality in older adults

To model COVID-19 mortality risk in older adults we fitted Cox proportional hazard regression models stratified by sex, including frailty penalties to accommodate multilevel data and random effects for the municipality of case occurrence, approximating iterations using the Newton-Raphson algorithm. Random effect estimates were exponentiated to calculate baseline mortality hazards across municipalities to represent geographical heterogeneity. Models were selected based on the Bayesian Information Criterion (BIC) and performance was assessed using Harrel’s c-statistic. To visualize increases in lethality risk for continuous variables and quantify uncertainty for estimations, we performed post-estimation simulations using bootstrapping with the *simPH* package. Proportional hazard assumptions were verified using Schoenfeld residuals.

#### Impact of age, structural determinants, and comorbidities on mortality risk

To investigate the role of age in predicting mortality compared to structural factors and comorbidities, we fitted sequential Cox proportional hazard models compared to only COVID-19 pneumonia as a predictor and introduced either age, structural factors (i.e. social lag index, private care), or increasing number of comorbidities as blocks. We compared models using changes in the Bayesian Information Criterion (BIC); the better models were those which either minimized or had large changes in BIC. We also separately analyzed outcomes and mortality for older adults without comorbidities to examine the role of structural factors compared to age in relation to disease severity and COVID-19 lethality. A p-value <0.05 was considered as the statistical significance threshold. All analyses were performed using R version 4.0.0.

## RESULTS

### COVID-19 in Mexican adults ≥60 years

As of May 9^th^, 2020, we observed 7,029 confirmed SARS-CoV-2 cases in adults aged ≥60 years, representing 21.0% of all confirmed COVID-19 cases (n= 33,460). Overall, the rate of positivity in this population has been 35.1% which represents a ratio of 1.43:1 in positivity amongst all evaluated cases. Despite the higher positivity rate, mortality rates are disproportionally higher in this population segment. Amongst all 3,353 confirmed lethal cases, 1,626 have been recorded in older adults, indicating significantly higher lethality (6.53% vs. 23.13%). When standardizing incidence rates to population estimates per each 1-year increase, the highest incidence rates occur between 55-65 years with a decrease in those aged >65 years; regardless, mortality remains steadily higher for adults aged ≥60 years (**Figure 1A, 1B**).

**FIGURE 1.**
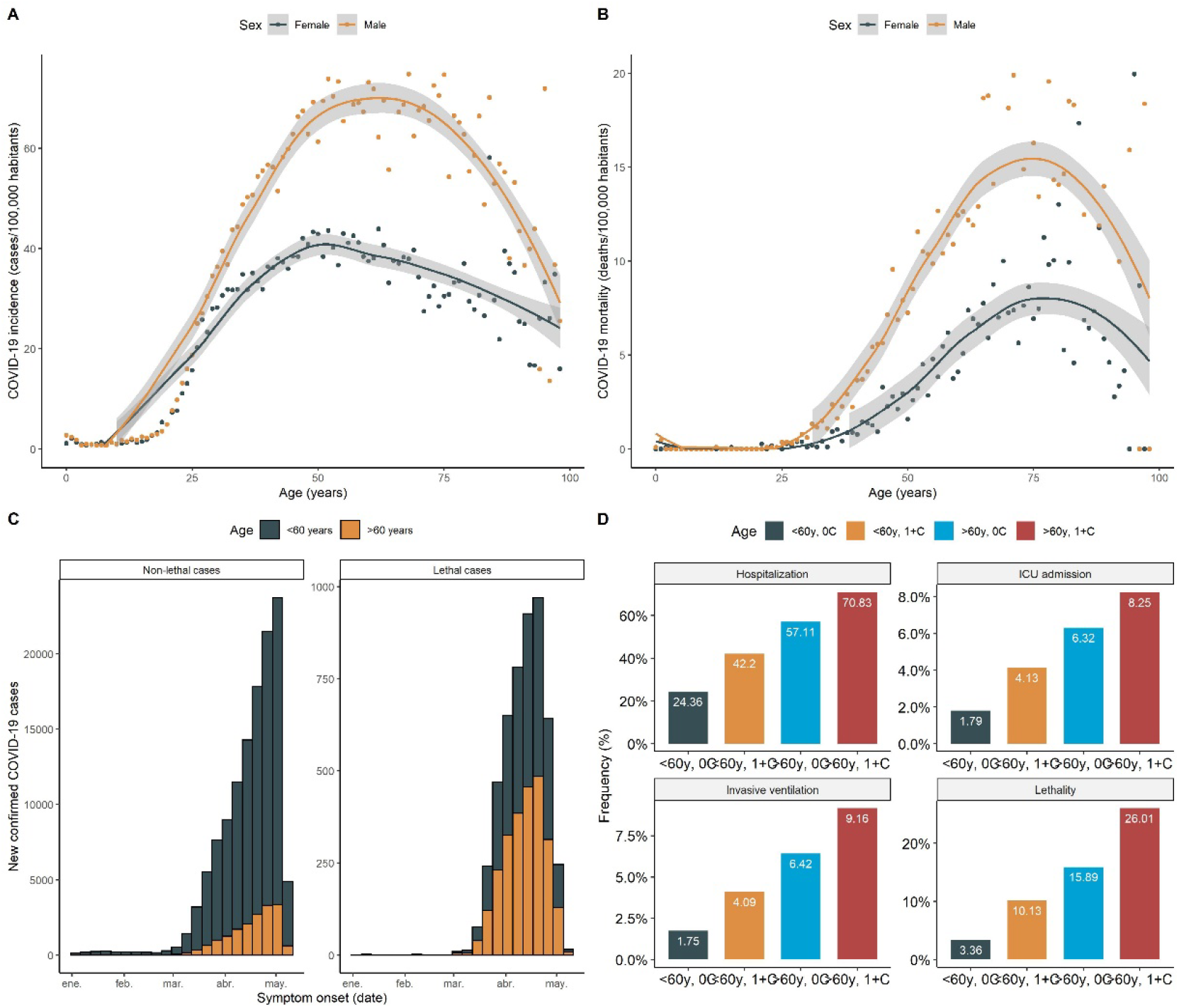
COVID-19 cases (A) and mortality rates (B) standardized according to age specific population projections for 2020 for each 1-year increment. The figure also displays the distribution of lethal and non-lethal cases stratified by age according to date of symptom onset (C) and distribution of COVID-19 outcomes comparing rates between those younger and older than 60 years (D).

Similarly, higher disease severity as assessed by hospitalization and ICU admission rates, as well as requirements for invasive ventilation, were more frequent in older adults. Furthermore, most comorbidities were clustered in older individuals and the rates of individuals having 2 or more comorbidities were markedly higher for older adults (25.5% vs. 12.7%, p<0.001, **Figure 1C, 1D**).

### COVID-19 outcomes and disease severity in older adults

We hypothesized that COVID-19 outcomes would be geographically heterogeneous; therefore, we fitted mixed effect models considering the municipality of case occurrence as a random effect. We identified that increasing age, male sex, smoking, diabetes, and obesity were associated with increased pneumonia risk; similarly, for hospitalization we identified an increased risk for older adults with CKD, COPD, diabetes, increasing age, and male sex. Notably, both pneumonia and hospitalization risk were higher for older adults living in municipalities with high social lag indexes, indicating an effect of structural inequalities besides the aforementioned comorbidities in further increasing COVID-19 severity (**Figure 2**). When assessing ICU admission risk factors, we identified increased likelihood with obesity (OR 1.37, 95%CI 1.07-1.74); similarly, the risk for invasive ventilation was higher for older adults with obesity (OR 1.45, 95%CI 1.16-1.81) and municipalities with high social lag index (OR 1.35, 95%CI 1.09-1.67). Notably, in older adults without comorbidities (n=1,900), age was not a significant predictor of pneumonia, the requirement for invasive ventilation, ICU or hospital admission. We only observed a significant role for age in predicting pneumonia when modeling an interaction effect with the social lag index (OR 1.034, 95%CI 1.002-1.067); moreover, the social lag index alone was associated with increased risk of hospitalization (OR 2.060, 95%CI 1.459-2.909) and requirements for invasive ventilation (OR 1.582, 95%CI 1.045-2.780).

**FIGURE 2.**
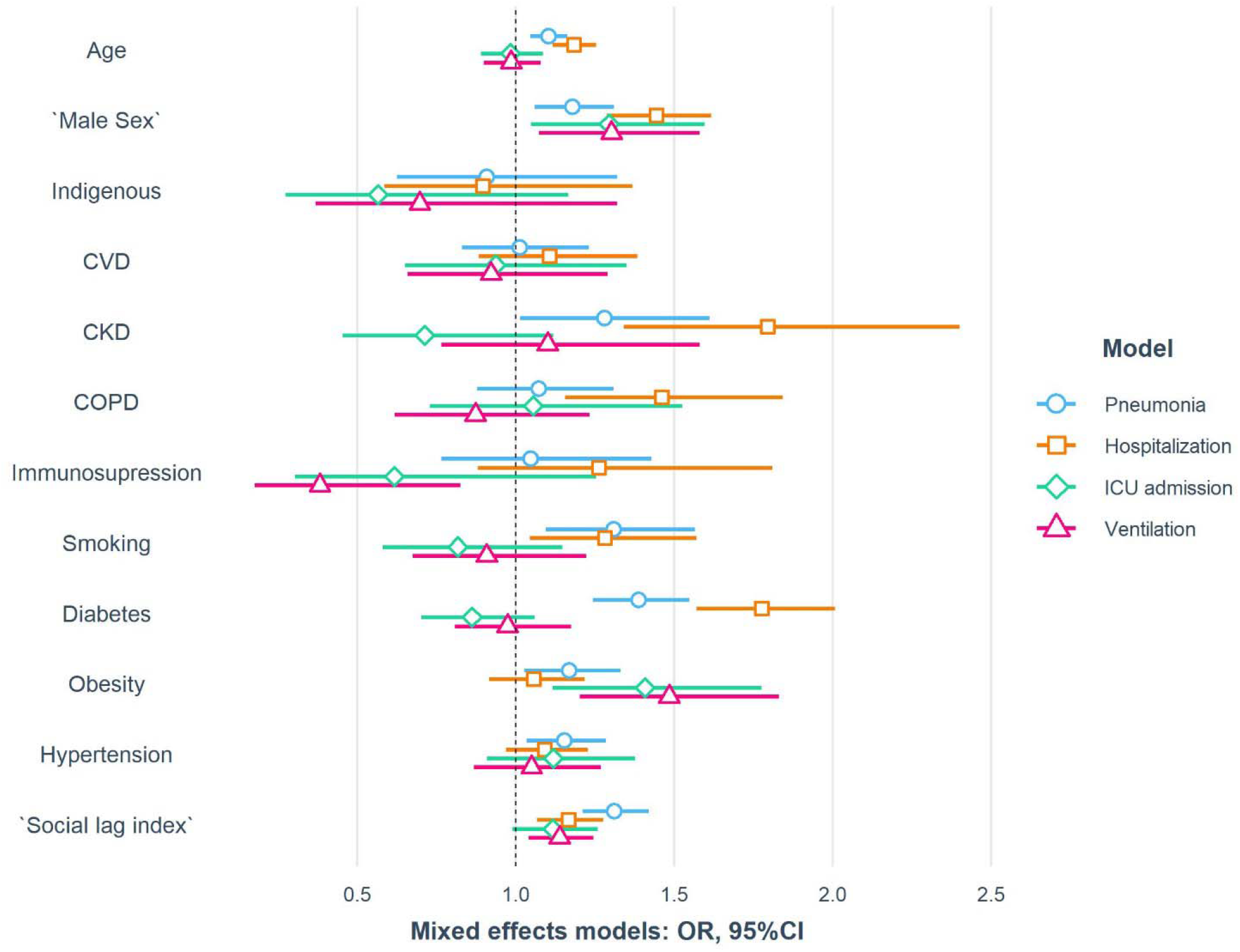
Risk factors for COVID-19 related pneumonia, hospitalization and intensive care unit (ICU) admission in older Mexican adults, modeled using logistic with mixed effects, including municipality of case occurrence as a random effect to control for regional inequalities.

### Health and structural determinants of COVID-19 mortality in older adults

Besides comorbidities, we identified that the social lag index was a predictor of COVID-19 lethality adjusted for age and sex (HR 1.327, 95%CI 1.174-1.500); the number of hospital beds per 1,000 individuals was a protective factor for mortality (HR 0.983, 95%CI 0.973-0.994) as was treatment received within a private healthcare facility (HR 0.403, 95%CI 0.288-0.563). In multivariable analyses, we identified a higher mortality risk associated with age, comorbid diabetes, obesity, immunosuppression, CKD, and pneumonia. Being treated in a public care facility and increased values of the social lag index were also predictors of COVID-19 lethality (**Figure 3A**).

**FIGURE 3.**
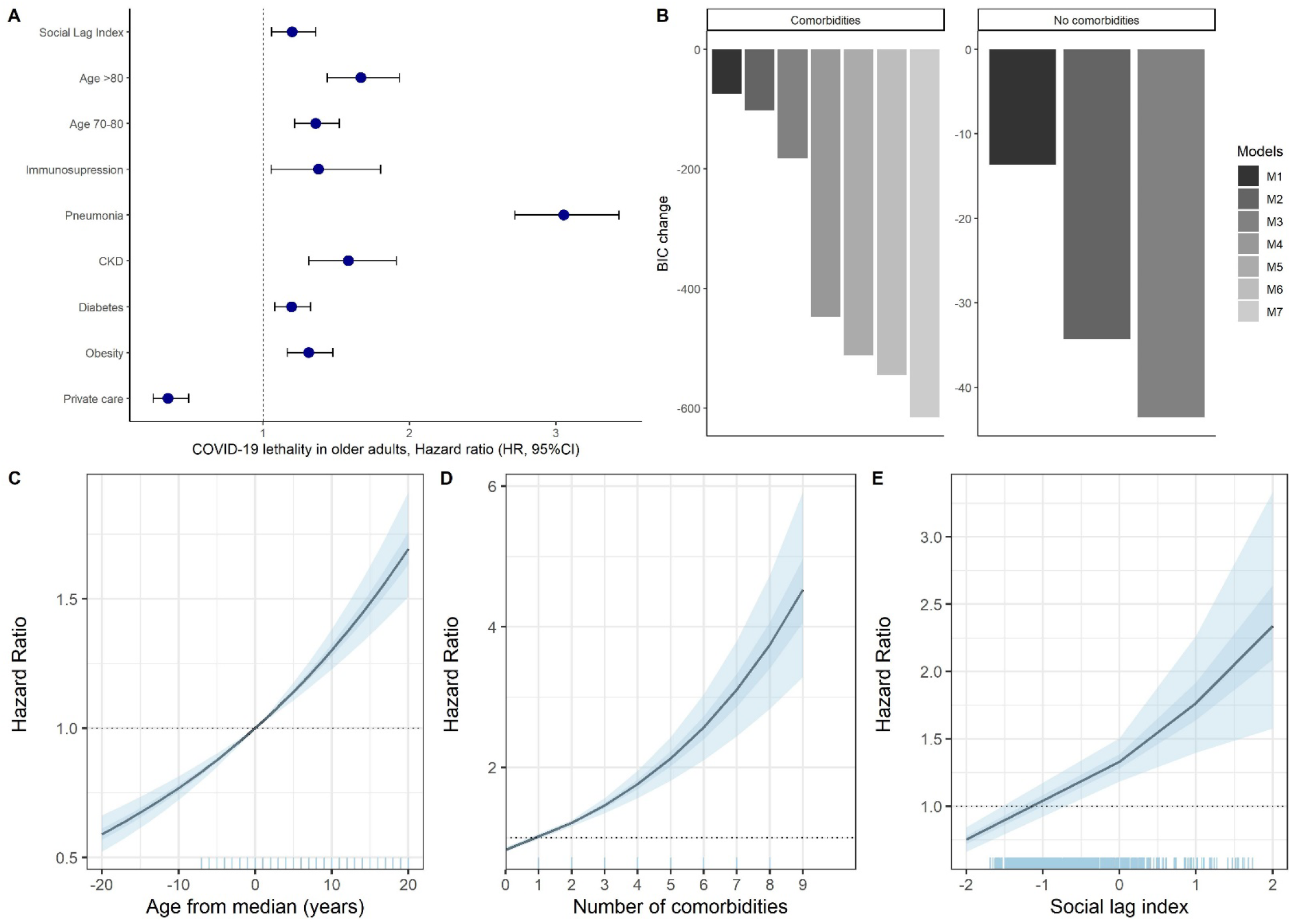
Risk factors for COVID-19 lethality in older Mexican adults, using Cox proportional Hazard regression models with frailty penalty to accommodate multilevel data (A). Panel **B** shows the changes in the Bayesian Information Criteria to estimate the role of age, structural factors and comorbidities in improving COVID-19 lethality risk predictions M0 is the reference model, which includes COVID-19 pneumonia, M1=M0+Age, M2=M0+Social lag index, M3=M0+Age+Social lag index, M4=M0+Comorbidities, M5=M0+Age+Comorbidities, M6=M0+Comorbidities+Social lag index, M7=M0+Age+Social lag index+Comorbidities comparing subjects with and without comorbidities. We also include post-estimation simulations to quantify the effect of covariates in increasing COVID-19 lethality risk for age compared to the median in older adults (72 years, C), an increasing number of comorbidities (D) and increases in the social lag index (E). COVID-19 mortality risk (HR)

#### Role of structural determinants, comorbidities, and age on COVID-19 severity and lethality

To assess improvements in model performance, we examined the effect of age, structural factors, and comorbidities in improving prediction of COVID-19 lethality. Overall, comorbidities offered the larger changes in predictive improvement for mortality, followed by structural factors; models that included age had markedly smaller changes in BIC. Similarly, the combination of comorbidities and structural factors showed lower BIC values compared to the combination of either of them with age (**Figure 3B**). The better model, which included age, structural factors, and comorbidities did not show marked improvements in predictive ability compared to the model which excluded age (c-statistics 0.728 vs. 0.722, respectively). To isolate the effect of age, we analyzed older adults without comorbidities; in these individuals, the social lag index was a risk factor (HR 1.338, 95%CI 1.083-1.652) and treatment in private care facilities a protective factor (HR 0.266, 95%CI 0.117-0.604) for mortality in addition to pneumonia (c-statistic=0.758). Addition of age, despite offering decreases in BIC, decreased the overall predictive capacity of the model (c-statistic=0.754). Overall, in individuals without comorbidities, structural factors were better predictors of mortality compared to age alone. Similarly, obesity remained as a significant predictor of mortality even in individuals without other comorbidities (HR 1.373, 95%CI 1.045-1.804) and was instead a predictor of increased risk of pneumonia (OR 1.948, 95%CI 1.473-2.576), hospitalization risk (OR 1.706, 95%CI 1.256-1.317) and requirement of invasive ventilation (OR 1.605, 95%CI 1.019-2.528), over chronological age. These latter findings suggest that the obesity paradox observed in other acute respiratory infections is not present in older individuals [16].

#### Post-estimation simulation of mortality risk prediction for continuous variables

When running post-estimation simulations in the model with all three categories, we identified that compared to median age amongst older adults (72 years), those younger than the median had lower mortality risk compared to those above. However, despite age being a predictor of mortality, we did not observe significantly higher risk in individuals aged >80 compared to individuals aged 70-80 years. Similarly, an increasing number of comorbidities and living in municipalities with increasing social lag index were also related to monotonic increases in mortality risk (**Figures 3C-E**). When considering baseline mortality hazard for older adults with COVID-19 across Mexican municipalities, we identified significant disparities; notably, higher risk areas were related to municipalities with higher social lag indexes which were mostly nearby metropolitan areas (**Figure 4A**). Furthermore, when plotting baseline hazard ratios using the number of hospital beds available per 1,000 inhabitants, we observed that higher risk occurred in municipalities with five or fewer beds per 1,000 inhabitants. Since the distribution of cases has not been homogeneous and most high-risk areas were near metropolitan areas, we performed a sub-analysis restricted to those locations (n=5377, 1122 deaths). Interestingly, the social lag index was not a significant predictor of mortality in metropolitan areas (HR 1.087, 95%CI 0.782-1.511), but being treated in a private care facility was (HR 0.489, 95%CI 0.331-0.725), suggesting a rural-urban dissociation on the factors which impact lethality.

**FIGURE 4.**
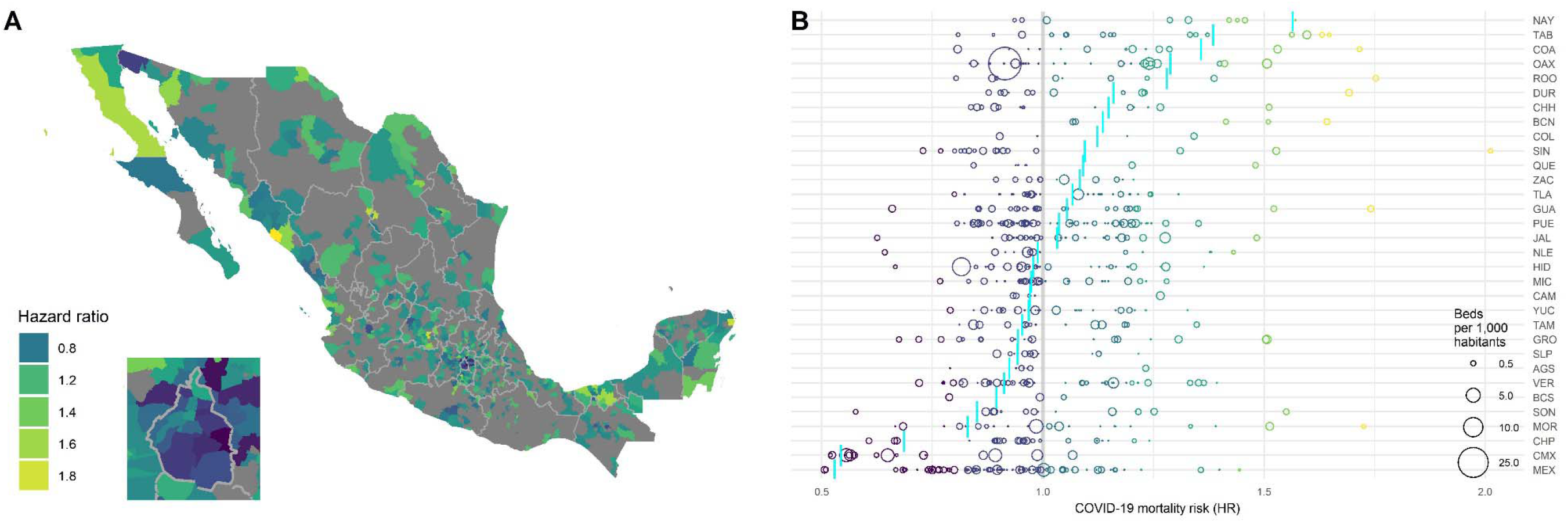
Geographical distribution of baseline COVID-19 lethality hazards across Mexican municipalities as modeled by mixed-effects Cox proportional risk regression models with frailty penalties (A). We also show Hazard of individual municipalities and the number of beds per 1,000 inhabitants in each municipality considering also a state-wide mortality hazard (blue line, B).

## DISCUSSION

Here, we demonstrate the role of health and structural socio-economic determinants in increasing COVID-19 lethality and disease severity independent of age in older Mexican adults. Notably, we observed that whilst age is a significant predictor of COVID-19 lethality, comorbidities, and structural health determinants likely play a larger role in increasing disease severity and conditioning risk of COVID-19 lethality in older patients. In older adults without comorbidities, age was not a predictor of disease severity with structural factors playing a larger role; in addition, mortality prediction in individuals without comorbidities did not significantly improve when considering age added to structural factors. We therefore confirmed a preoccupying association of increased COVID-19 lethality with increasing values of the social lag index, which considers deficiencies or unequal access to education, healthcare, living conditions, and basic services; furthermore, being treated in private facilities also decreases the risk of COVID-19 lethality, suggesting a significant gap between care received in public compared to private facilities for older adults or better accessibility for private care. These results demonstrate that the consequences of large socio-economic inequalities within Mexico, which represent added burdens to multimorbidity in older adults, both of which are better predictors of mortality compared to chronological age, and must be addressed as the pandemic transitions to more rural areas where limited access to health services and older population structures with adverse multimorbidity profiles increase mortality risk.

Data from countries affected by the COVID-19 pandemic have reported higher mortality rates in older adults, particularly in those with an increasing number of comorbidities [17]. These observations have led to distinct problems concerning healthcare decisions in the face of COVID-19. Given the overwhelming effect of COVID-19 on healthcare burden for countries with older population structures, many systems have opted for health resources to be allocated based mainly on chronological age [18,19]. Whilst evidence of an age-dependent increase in the expression of the *ACE2* gene has been reported and an increasing number of comorbidities are readily observed in older adults with COVID-19, data which demonstrates biological effects of chronological aging over multimorbidity and health disparities on outcomes has not been reported [5,20]. Instead, additional effort should be carried out to reduce disparities in healthcare access, reduce socio-economic inequalities and improve care in older adults with multimorbidity to reduce its effect on increasing COVID-19 severity, given their predominant effects over chronological age in increasing COVID-19 severity and lethality [21,22]. A previous analysis by our group which considered COVID-19 cases in Mexico highlighted how the cardio-metabolic burden in younger patients approximated mortality risk to those observed in older COVID-19 patients without comorbidity [23]. Our results in this present study show that chronological age only offer modest improvements in mortality risk predicction and does not predict disease severity in older adults without comorbidities. This evidence advocates for bioethical decisions on resource allocation in overwhelmed healthcare systems to not be based on chronogical age as an exclusive criteria, but rather to integrally consider comorbidity profiles and establish a framework which works towardws ameliorating the impact of socio-economic inequalities on COVID-19 outcomes for older adults as the pandemic progresses.

The response of Mexican authorities to promote social distancing amongst older adults has thus far been consistent. A nation-wide stipend was established by the current administrtation to support older adults throughout Mexico and advanced payments were released to reduce the financial burden of COVID-19 [24]. Despite these policies, our data shows that older adults who live in highly marginalized municipalities remain at higher risk of COVID-19 severity and mortality, which might likely indicate unaddressed gaps that must be considered to reduce the impact of COVID-19 on older adults, particularly from highly marginalized communities and ethnic groups within Mexico [25]. Inequalities which might arise once social distancing is lifted must also be addressed to facilitate reincorporation of older adults into daily life and geriatric care must be secured to reduce the impact of social distancing on the general health of older adults, particularly those living in isolation and with multimorbidity [26].

Obstacles for healthcare access in older adults have been consistently associated with worsened outcomes and increased all-cause mortality, particularly for those facing social inequalities [27]. Previous research has shown that negative self-perceived aging influences healthcare-seeking behavior, particularly in settings of limited healthcare access [28,29]. Our results suggest that reduced healthcare access, as measured by increases in the social lag index, and lesser number of available hospital beds are associated with increased COVID-19 severity and lethality in older adults, likely reflecting the effect of structural inequalities across Mexican municipalities. When exploring the interaction of this index with increasing age, we observed a non-significant increased trend for mortality risk (HR 1.006, 95%CI 0.996-1.016, p=0.053), suggesting that these deficits may have larger impacts in the oldest old. A recent geroscience approach to the COVID-19 pandemic in the US, Italy, and China demonstrated an exponential increase in COVID-19 lethality with age and similarly observed male-biased mortality as in the Mexican data, potentially implicating aging mechanisms into COVID-19 lethality in older adults [30]. However, as shown by our results, in older adults without comorbidities the effect of age was not a predictor of disease severity; furthermore, structural factors and comorbidities were better predictors of mortality compared to chronological age. These findings of individual and structural factors relating the individual with its environment are in line with the multidimensional concept encapsulated by intrinsic capacity, which is a composite of physical and mental capacities which define and individual’s functional ability. These deficits might be responsible for the increased COVID-19 risk observed in older adults. Intrinsic capacity should be evaluated as a risk factor for COVID-19 and other infectious diseases in older adults in future studies to better understand it as a predictor of mortality over chronological aging [31,32].

Ageism and self-perceived aging might impact healthcare-seeking decisions in older adults, particularly under a widespread perception of increased healthcare burden during the COVID-19 pandemic [11,33]. An additional factor to consider in this setting is the fact that awareness of COVID-19 symptoms is directed towards typical disease presentations and might not account for atypical symptoms and presentations which are more likely to occur in older patients [34,35]. Overall, given the significant impact of COVID-19 on older adults, our findings call for more integral approaches to spread awareness of COVID-19 in older adults in Mexico and other countries, addressing information gaps which might reduce healthcare-seeking behavior and reducing ageism as a criteria for bioethical decision-making. Moreover, evidence regarding atypical COVID-19 presentations and identifying factors that decrease mortality in older adults to increase awareness and promote healthcare-seeking behavior in high-risk remain as a relevant unaddressed gap which must be undertaken by future studies Our approach is robust given that it considers national data on all confirmed COVID-19 cases in older adults with significant representation of most regions within Mexico. Inclusion of multilevel data allowed us to identify structural factors related to COVID-19 lethality and outcomes and to contrast the role of these factors and comorbidities as predictors of COVID-19 severity and mortality, highlighting areas of opportunity to address these and future affections which will likely also impact older adults. A potential limitation of our study is the lack of symptom-specific data which could be helpful to characterize atypical presentations of COVID-19 in older adults and identify early predictors of disease severity and potential recovery as has been seen in some series, and which remains to be addressed in future studies to better characterize the course of COVID-19 in older adults [36]. Finally, an additional limitation of our work could be related to underreporting of mild cases in marginalized areas due to a lack of widespread testing, which might inflate mortality estimates for those regions, but which also would indicate a significant challenge for handling the COVID-19 epidemic in marginalized communities moving forward. Our findings could inform public policy related to ethical decision making for older adults by prompting consideration of factors beyond chronological age and promote further studies in older populations, which remains imperative to address aging in populations undergoing an epidemiological transition with increased morbidity burden, such as Mexico.

In conclusion, we characterized predictors of COVID-19 outcomes in older Mexican adults, including potential predictors related to structural inequalities which increase COVID-19 severity and lethality risk. We demonstrated that comorbidity profiles and structural factors are better predictors of COVID-19 severity and mortality compared to chronological age. Overall, baseline COVID-19 mortality hazards are unequally distributed for older adults in Mexico. Special attention should be given to underprivileged older populations to ameliorate the impact of these factors which increase lethality in older adults with multimorbidity. These findings could aid in shifting the current clinical and ethical decision-making focus away from chronological aging and onto a more integral and comprehensive geriatric care approaches.

## Data Availability

All data sources and R code are available for reproducibility of results at https://github.com/oyaxbell/covid_aging_mx

## ACKNOWLEDGMENTS

NEAV, JPBL, AVV, AMS, and CAFM are enrolled at the PECEM program of the Faculty of Medicine at UNAM. JPBL and AVV are supported by CONACyT. The authors would like to acknowledge the invaluable work of all of Mexico’s healthcare community in managing the COVID-19 epidemic. Its participation in the COVID-19 surveillance program has made this work a reality, we are thankful for your effort.

## AUTHOR CONTRIBUTIONS

Research idea and study design OYBC, AGD, LMGR; data acquisition: OYBC, AGD; data analysis/interpretation: OYBC, JPBL, NEAV, AVV, AGD, CGP, CAAS, LMGR; statistical analysis: OYBC, AGD; manuscript drafting: OYBC, AGD, NEAV, JPBL, AMS, CAFM; supervision or mentorship: OYBC, LMGR. Each author contributed important intellectual content during manuscript drafting or revision and accepts accountability for the overall work by ensuring that questions about the accuracy or integrity of any portion of the work are appropriately investigated and resolved.

---

**Table 1.**
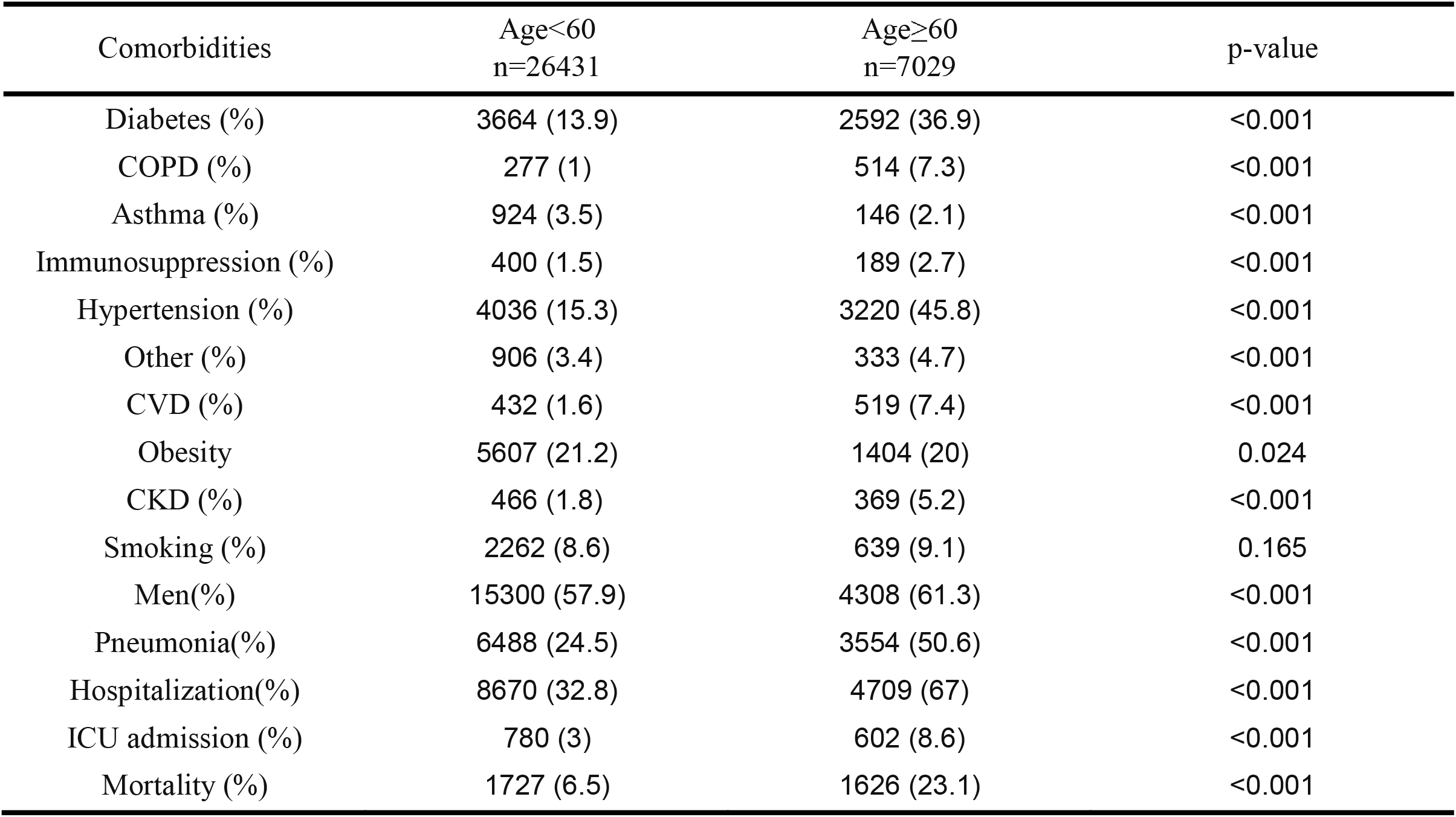
Descriptive statistics positive SARS-CoV-2 cases in Mexico at 09/05/2020. *Abbreviations:* ICU= intense care unit; COPD= chronic obstructive pulmonary disease; CKD= chronic kidney disease; CVD= cardiovascular disease.

